# Protocol for the development and validation of machine-learning models for predicting the risk of hypertriglyceridemia in critically ill patients receiving propofol sedation using retrospective data

**DOI:** 10.1101/2024.08.17.24312159

**Authors:** Jiawen Deng, Kiyan Heybati, Hemang Yadav

## Abstract

**Introduction:** Propofol is a widely used sedative-hypnotic agent for critically-ill patients requiring invasive mechanical ventilation (IMV). Despite its clinical benefits, propofol is associated with increased risks of hypertriglyceridemia. Early identification of patients at risk for propofol-associated hypertriglyceridemia is crucial for optimizing sedation strategies and preventing adverse outcomes. Machine learning (ML) models offer a promising approach for predicting individualized patient risks of propofol-associated hypertriglyceridemia.

**Methods and analysis:** We propose the development of a ML model aimed at predicting the risk of propofol-associated hypertriglyceridemia in ICU patients receiving IMV. The study will utilize retrospective data from four Mayo Clinic sites. Nested cross-validation (CV) will be employed, with a 10-fold inner CV loop for model tuning and selection as well as an outer loop using leave-one-site-out CV for external validation. Feature selection will be conducted using Boruta and LASSO-penalized logistic regression. Data preprocessing steps include missing data imputation, feature scaling, and dimensionality reduction techniques. Six ML algorithms will be tuned and evaluated. Bayesian optimization will be used for hyperparameter selection. Global model explainability will be assessed using permutation importance, and local model explainability will be assessed using SHapley Additive exPlanations (SHAP).

**Ethics and dissemination:** The proposed ML model aims to provide a reliable and interpretable tool for clinicians to predict the risk of propofol-associated hypertriglyceridemia in ICU patients. The final model will be deployed in a web-based clinical risk calculator. The model development process and performance measures obtained during nested cross-validation will be described in a study publication to be disseminated in a peer-reviewed journal. The proposed study has received ethics approval from the Mayo Clinic Institutional Review Board (IRB #23-007416).

**Strengths and limitations of this study:**

- Robust external validation using a nested cross-validation (CV) framework will help assess the generalizability of models produced from the modeling pipeline across different hospital settings.
- A diverse set of machine learning (ML) algorithms and advanced hyperparameter tuning techniques will be employed to identify the most optimal model configuration.
- Integration of feature explainability will enhance the clinical applicability of the ML models by providing transparency in predictions, which can improve clinician trust and encourage adoption.
- Reliance on retrospective data may introduce biases due to inconsistent or erroneous data collection, and the computational intensity of the validation approach may limit replication and future model expansion in resource-constrained settings.

## INTRODUCTION

Propofol is a sedative-hypnotic agent commonly used for sedation in critically ill adults requiring invasive mechanical ventilation (IMV) [1]. It is recommended as one of the first-line regimens for this indication by the 2018 Pain, Agitation/sedation, Delirium, Immobility, and Sleep (PADIS) guidelines [2] due to its rapid onset and short duration of action. As a highly lipophilic drug, propofol is formulated in a 10% fat emulsion, typically using soybean oil [1]. However, this formulation has the disadvantage of predisposing patients to hypertriglyceridemia [1,3]. Up to 10% of patients who develop propofol-associated hypertriglyceridemia may progress to pancreatitis [4], which substantially increases these patients’ risks of morbidity and mortality in the ICU [5].

Thus, it is crucial to assess potential risk factors associated with the development of propofol-associated hypertriglyceridemia when selecting sedative regimens. Several retrospective cohort studies have identified important predictors of hypertriglyceridemia following propofol sedation, including advanced age [4], propofol dose and duration, body mass index (BMI), illness severity, and concomitant medications [6,7]. However, the question of how this knowledge can be applied systematically for guiding clinical practice remains unanswered.

Machine-learning (ML), which is a modeling paradigm that can identify complex and non-linear patterns in large datasets [8,9], has proven effective for providing individualized patient risk stratification in medical diagnoses and prognoses [10]. ML models could potentially help identify recently intubated ICU patients who are at higher risks for developing propofol-associated hypertriglyceridemia, allowing for these patients to be switched to alternative sedation regimens. Thus, the objective of the proposed ML development study is to create a ML-powered clinical calculator to aid in selecting sedating regimens based on personalized patient risk predictions of developing propofol-associated hypertriglyceridemia in ICU settings.

## METHODS AND ANALYSIS

The proposed study will be conducted and reported in accordance with the Transparent Reporting of a Multivariable Prediction Model for Individual Prognosis or Diagnosis+AI (TRIPOD+AI) Checklist for Prediction Model Development and Evaluation [11].

### Data sources

The proposed study involves a secondary analysis of data from a multi-centered retrospective cohort investigation conducted at Mayo Clinic sites in the United States. The original retrospective study included consecutive adults (≥18 years of age) admitted to one of 11 ICUs across four Mayo Clinic sites: 1) Mayo Clinic Rochester, 2) Mayo Clinic Phoenix, 3) Mayo Clinic Jacksonville, and 4) Mayo Clinic Health System (MCHS) community sites in Mankato, Minnesota and Eau Claire, Wisconsin. Patient inclusion criteria for the proposed model development study is the same as the eligibility criteria in the original retrospective study, which included: 1) admission to one of the study ICUs between May 5^th^ 2018, and June 30^th^ 2023, 2) required IMV for greater than 24 hours, and 3) received continuous propofol infusion for >24 hours. Exclusion criteria included: 1) development of hypertriglyceridemia (defined as serum triglyceride levels >400 mg/dL) [5] prior to propofol infusion, and 2) lack of prior authorization for medical records to be assessed for research purposes. The index ICU admission for eligible patients were identified through institutional data warehouses (Mayo Clinic ICU DataMart and Unified Data Platform).

### Outcomes of interest

The goal of model development is to predict the probability of hypertriglyceridemia within 10 days following the start of propofol infusion. Hypertriglyceridemia is defined as serum triglyceride levels exceeding 400 mg/dL [5]. The predicted probability estimates will be transformed into a binary classification to categorize patients into high risk and low risk groups for developing hypertriglyceridemia based on a decision threshold. The anticipated usage setting for the model is during the ICU admission process or shortly before/after intubation and sedation for IMV.

### Nested cross-validation

A nested cross-validation (CV) methodology [12] will be used to evaluate the performance and consistency of the overall ML modeling process. Nested CV involves two layers of CV (**Figure 1**):

**Figure 1:**
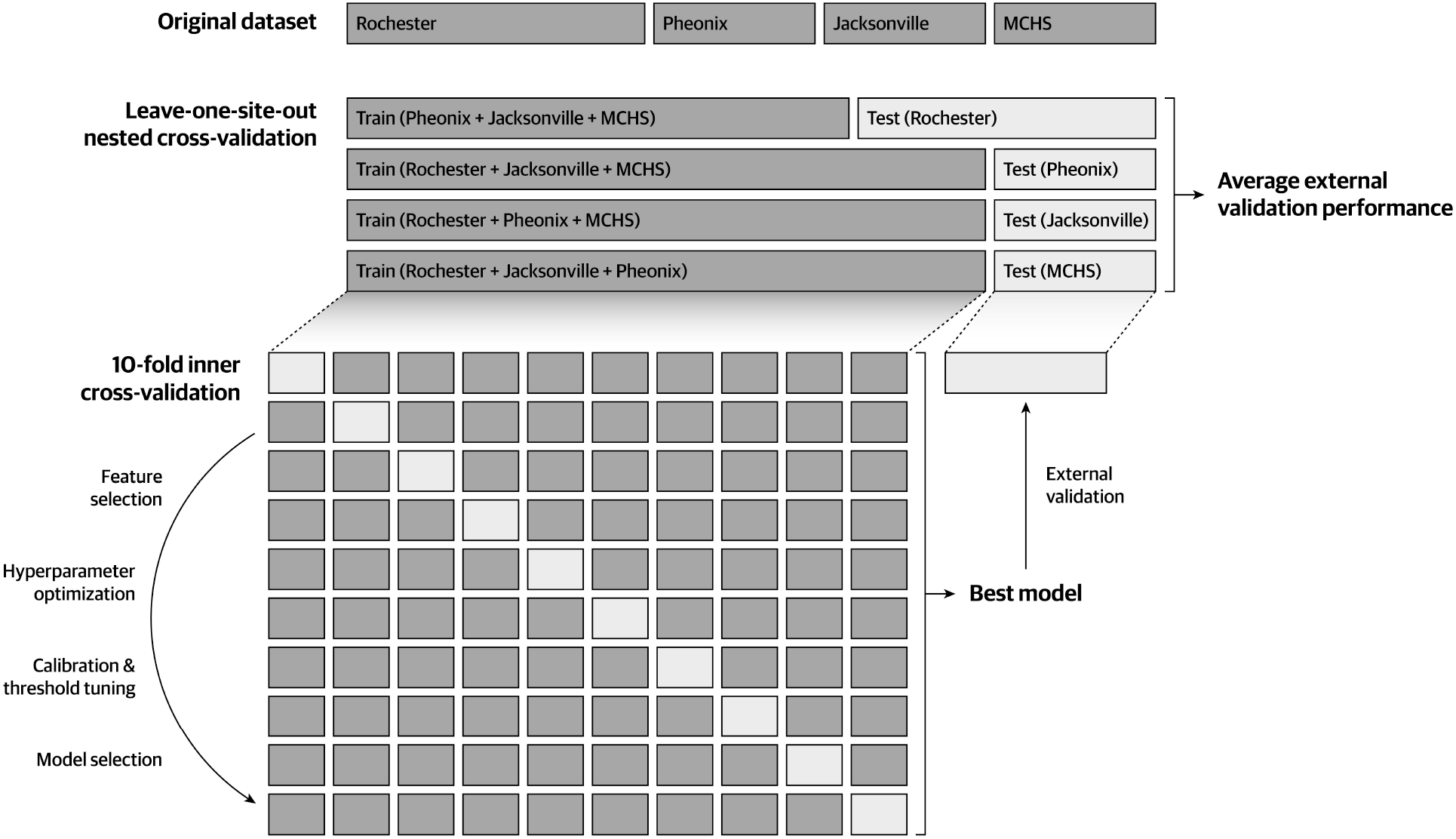
Illustration of the proposed nested cross-validation approach. The outer loop consists of a leave-one-site-out cross-validation approach for repeated external validation of models produced by the inner loop. The inner loop involves a 10-fold cross-validation approach for model tuning and selection. **Abbreviations: MCHS** Mayo Clinic Health System (community sites).

1. **Inner CV loop:** The inner CV loop is used to tune each ML algorithm and to select the best-performing model. This is similar to regular, non-nested “flat” CV.
2. **Outer CV loop:** The dataset is divided into *k* outer folds. In each iteration of the outer loop, one fold is held out as the test set while the remaining *k-1* folds are used for model tuning and selection using the inner CV loop. The main function of the outer loop is to estimate the performance of the best-performing model selected by the inner loop [13].

For our proposed study, we will use stratified 10-fold CV as our inner CV loop for hyperparameter tuning and model selection. For the outer CV loop, we will use leave-one-site-out CV (LOSO-CV). In our study, LOSO-CV involves tuning, selecting, and training models on data from three of the four included Mayo Clinic sites and externally validating the best performing model on data from the remaining site. This is repeated four times so that each Mayo Clinic site serves as the external validation set at least once. In essence, we are externally validating our modeling process four times to better assess how our models will perform on new, unseen data [14]. Following the nested CV process, we will run the inner CV loop on the entire dataset to generate the final production model for deployment into a clinical calculator.

### Feature selection and sample size assessment

Candidate features will be first filtered based on availability (≤10% of missing data) and expert domain (see **Table 1** for the list of candidate features). For each training set in the outer CV loop, the dimension of the feature set will be further reduced using random-forest-based Boruta [15] and Least Absolute Shrinkage and Selection Operator (LASSO) penalized logistic regression [16]. During feature selection, the regularization hyperparameter α of the LASSO penalized logistic regression model will be determined via a grid-search of 1000 α values along the regularization path to minimize the LASSO objective function across 10-fold CV. A union of features selected by the two methods will be chosen as the final feature set used for predictive modeling.

**Table 1:**
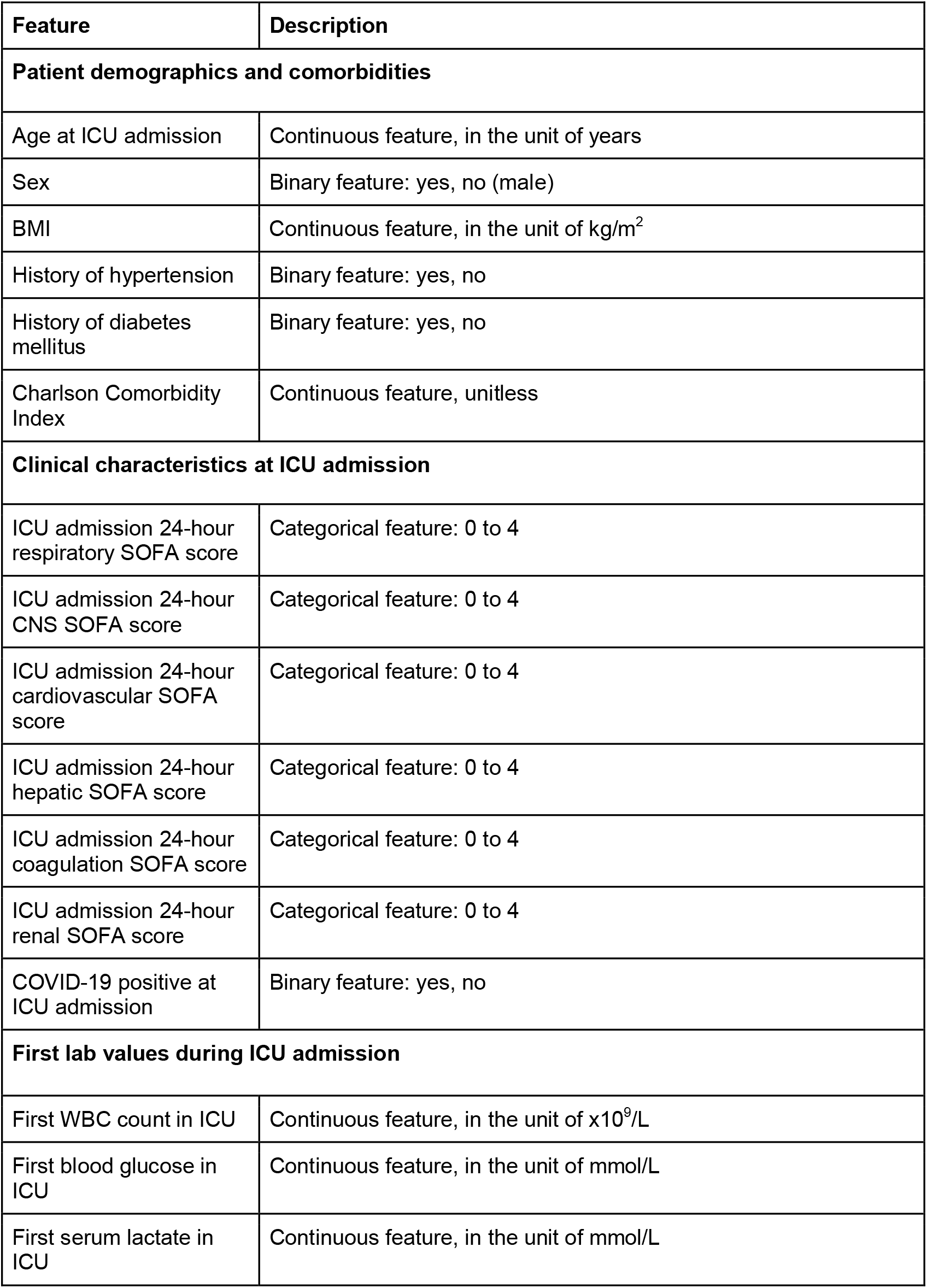
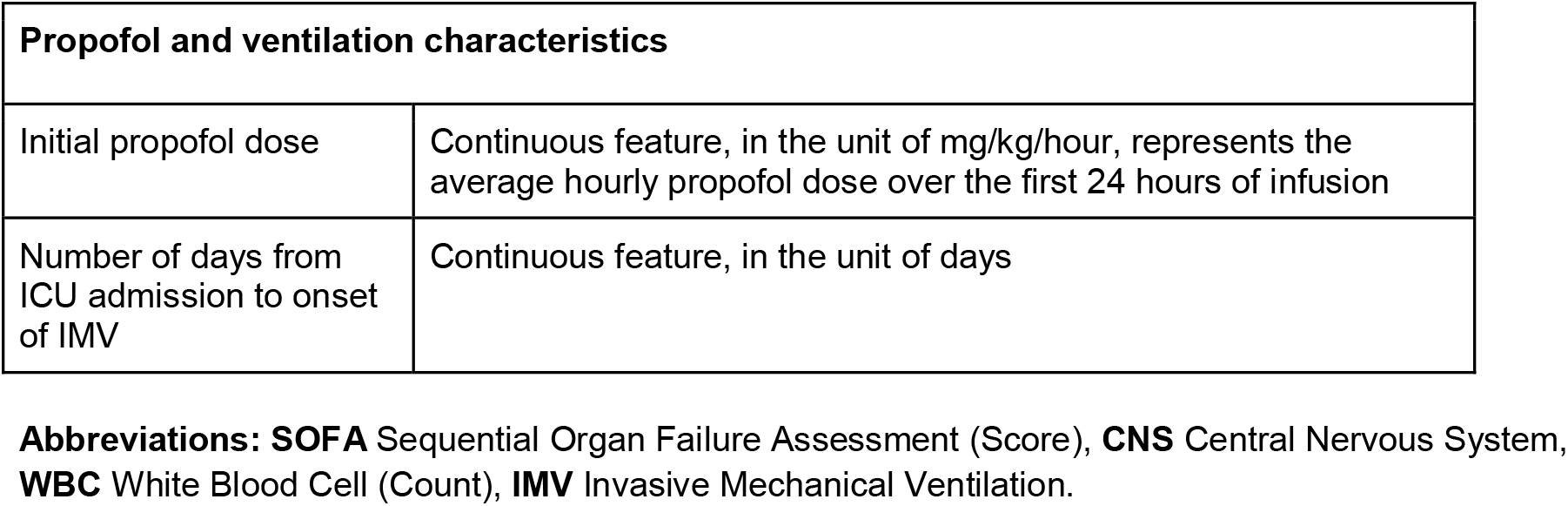
Candidate features selected based on data availability and expert domain.

The goal of the feature selection process is to follow the “one-in-ten” rule—which states that there should be 10 patients with the target outcome in the dataset to support every one feature included to ensure stable ML model predictions [17,18]—as closely as possible. Given that our previous analyses involving our dataset showed that there were 851 patients who developed hypertriglyceridemia following propofol administration (without applying the 10 day post-propofol-initiation restriction) [19], we will likely have capacity for 70-80 features. This is far greater than the number of candidate features that we expect to include in the model, thus we anticipate that our dataset will contain enough case samples for modeling purposes.

### Data preprocessing

Within each iteration of the outer CV loop, the training set will be split into 10 folds of different training and testing subsets for the inner CV loop. A data preprocessing pipeline will be constructed to identify and impute missing data, correct for class imbalance, and improve compatibility with ML algorithms before the data is entered into the ML models (**Figure 2**).

**Figure 2:**
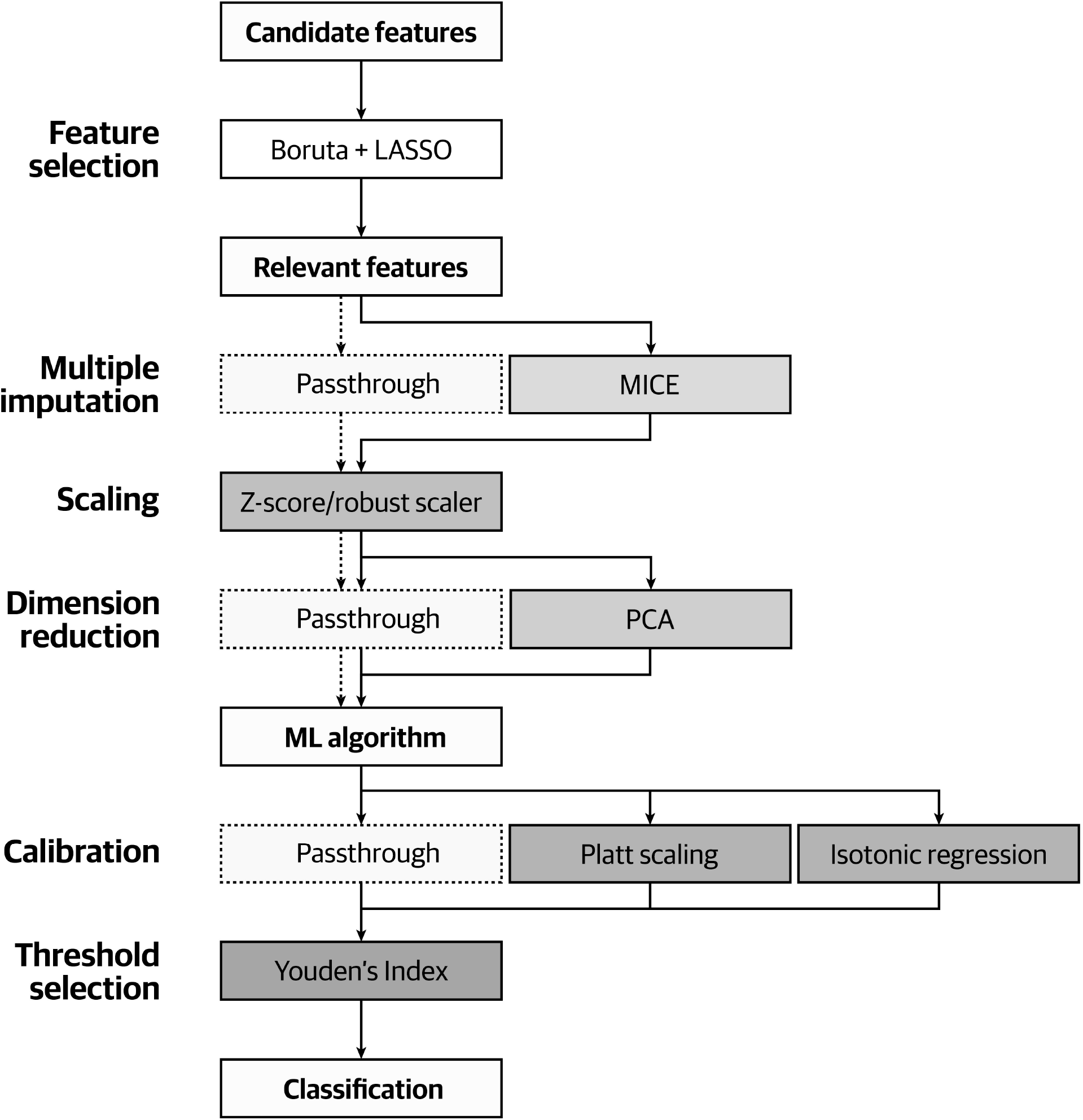
Flowchart illustrating the flow of training data through our proposed modeling pipeline. Because principal component analysis requires a complete dataset with no missing data, model configurations that did not use imputation (and relied on machine-learning algorithms’ native missing data handling methods) cannot use principal component analysis. Flow of data through configurations without imputation is shown using the dotted arrows. **Abbreviations: LASSO** Least Absolute Shrinkage and Selection Operator (penalized logistic regression), **MICE** Multivariate Imputation by Chained Equations, **PCA** Principal Component Analysis, **ML** Machine-Learning.

#### Categorical data encoding

As all of the candidate categorical features are non-ordinal, they will be transformed via one-hot encoding into multiple binary features [20].

#### Missing data imputation

Because we expect the amount of missing data in the dataset to be low (≤10%), we will assume missing data to be missing at random and perform missing data imputation. Imputation will be conducted using Multivariate Imputation by Chained Equations (MICE) [21]. As previous research had suggested that the number of MICE iterations should be determined based on the highest percentage of data missing (e.g., if 30% of data needs to be imputed for one feature, then MICE will be run for 30 iterations) [22,23], we will run MICE for 10 iterations to account for the maximum missing data percentage of 10%. The training and testing subsets will be imputed separately to avoid data leakage [24]. When assessing and tuning ML algorithms with native missing data handling methods, we will treat the use of MICE as a hyperparameter to trial both MICE and the algorithms’ native missing data handling approaches.

#### Data imbalance and resampling

Given that we aim to develop a model that can perform probability predictions, we will not resample the dataset even if the dataset is imbalanced. However, we will tune ML algorithms’ weight scaling factors as a hyperparameter during model tuning to help reduce bias towards the majority class.

#### Feature scaling

Before the dataset is entered into dimensionality reduction steps and ML algorithms, the normality of each continuous feature within the dataset will be assessed using the Shapiro–Wilk test [25] and by visual inspection of the features’ histograms [26]. If the distribution of all continuous features are determined to be close to normal, the continuous features will be Z-score normalized to center the feature data around the mean and scale the data according to its standard deviation [27]. If any of the continuous features were not close to normal, the continuous features will be transformed using robust normalization, which centers the data around the median and scales the data around its interquartile range [27]. Compared to Z-score normalization, robust normalization is more robust towards the presence of outlier values in the dataset [28].

#### Dimensionality reduction

Potential multicollinearity in the final feature set will be assessed using variance inflation factors (VIFs) [29]. If the VIF for any feature exceeds 5, we will trial the use of principal component analysis (PCA) as a dimensionality and multicollinearity reduction approach in our data preprocessing pipeline. The lowest number of principal components needed to explain at least 95% of the variance will be kept [30].

### Hyperparameter tuning

Within each iteration of the outer CV loop, we will tune and evaluate:

1. Two classical ML algorithms: 1) logistic regression (with LASSO, ridge, or elastic net penalization) and 2) support vector machines (SVMs, with linear, polynomial, radial basis function, or sigmoid kernels).
2. Three ensemble ML algorithms: 1) random decision forest, 2) Light Gradient-Boosting Machine [31] (LightGBM, with either regular Gradient Boosting Decision Trees [GBDT] algorithm [32] or Dropouts meet Multiple Additive Regression Trees [DART] algorithm [33]), and 3) eXtreme Gradient Boosting (XGBoost, with either regular gradient boosting decision trees [GBDT] algorithm [32] or Dropouts meet Multiple Additive Regression Trees [DART] algorithm [33]) [34].
3. A multilayer perceptron (MLP) neural network with 2, 3, or 4 hidden layers. Each hidden layer will use a rectified linear unit (ReLU) activation function with Kaiming kernel initialization [35]. Each ReLU layer will be followed by a dropout layer for regularization [36] and a batch normalization layer to improve training stability and speed [37]. All hidden layers will use the same number of neurons determined via hyperparameter tuning. The output layer will use a sigmoid function to produce the predicted probability. An AdamW optimizer will be used [38]. An example of the proposed network architecture with 2 hidden layers is shown in **Figure 3**.

**Figure 3:**
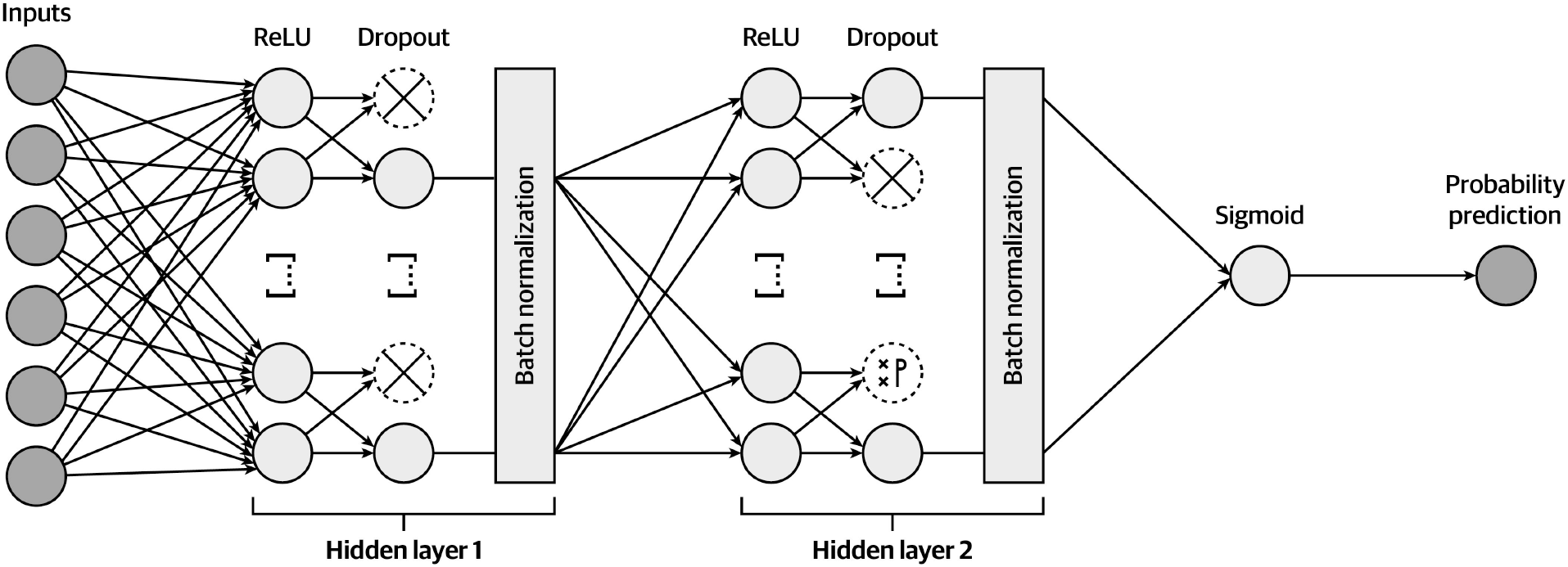
Example schema of the proposed network architecture, with dropout and batch normalization components in each hidden layer. An example with 2 hidden layers is shown. **Abbreviations: ReLU** Rectified Linear Unit.

Each algorithm will be tuned to minimize cross-entropy loss across stratified 10-fold CV. The full list of hyperparameters that will be tuned for each model is tabulated in **Supplementary Table S1 to S17**. The optimal hyperparameters will be selected using Bayesian optimization [39].

Bayesian optimization starts with several initial rounds of random hyperparameter searches to gather data points for building a probabilistic model that predicts the performance of different hyperparameter combinations. An acquisition function then uses this model to identify the most promising hyperparameter combinations for the next round of evaluations. The results are used to update the probabilistic model, and the process is repeated until a pre-established performance budget is exhausted [40]. In essence, Bayesian optimization can be thought of as a more intelligent and “guided” version of random hyperparameter searching, and it is empirically considered to outperform traditional grid-search and random search approaches [41]. Because random search can reliably identify hyperparameter combinations from the top 5% of the most performant combinations with 60 iterations [39], we aim to perform Bayesian optimization at or beyond this performance budget. With computational limitations, we estimate that Bayesian optimization will be performed for at least 200 iterations per model.

### Calibration

As the objective of our hyperparameter tuning process is to minimize cross-entropy loss, we can reasonably assume that our models will be well-calibrated. Nevertheless, we will trial Platt/sigmoidal scaling [42,43], isotonic regression [42], or no further calibration to assess if our calibration performance can be further improved. The calibration approach with the lowest average cross-entropy loss on stratified 10-fold cross-validation will be selected.

### Threshold selection

Following calibration, each model will undergo threshold tuning to maximize their binary classification performance. Youden’s Index [44] will be calculated for every classification threshold between 0.01 and 0.99 at an interval of 0.01, and the threshold with the highest average Youden’s Index across stratified 10-fold cross-validation will be selected as the most optimal threshold.

### Model selection and evaluation

We will follow a previously published framework proposed for the evaluation of clinical prediction models [45] to perform model selection and external validation. The classification performance of each tuned and calibrated model will be assessed using the area under the curve of the receiver operating characteristic curve (AUC-ROC), accuracy, sensitivity, specificity, and Youden’s Index. Calibration performance will be assessed using Brier score.

During model selection within the training set of each outer CV loop, all metrics will be produced and averaged across 10-fold cross-validation, and 95% confidence intervals (CIs) will be used to assess performance variance. The model configuration with the best classification and calibration performance will be selected as the most optimal configuration.

For external validation within each iteration of the outer CV loop, the same set of classification and calibration metrics will be generated using the most optimal model configuration and data from the held-out site not involved in model tuning and selection. The external validation metrics will be averaged across all 4 iterations of the outer CV loop to assess the generalization performance of our modeling process, and 95% CI will be used to assess the variance of the generalization performance.

### Model explainability and fairness

To understand how the final production model makes its predictions and to assess model bias arising from over-reliance on patient demographics, we will use the permutation importance method to assess which features have the greatest impact on the performance of our final production model [46]. Permutation importance is a global explainability method that involves repeatedly shuffling individual predictors in the training dataset, which effectively renders the predictor useless to the model. It then observes how this operation affects model performance, thereby identifying the most influential features. To understand how the production model makes individual predictions, we will use permutation-based SHapley Additive exPlanations (SHAP) [47], which is a local explainability method that produces an estimate of the direction and magnitude of each features’ effect on individual predictions. A bee swarm summary plot will be used to illustrate SHAP feature attributions for each patient in our dataset.

### Model deployment

The final production model will be integrated into a web-based clinical risk calculator based on *shiny*. SHAP waterfall plots will be provided for each individual prediction generated through the calculator to allow users to understand how the model is generating its predictions and to assess whether the predictions make clinical and biological sense.

Implementation of permutation SHAP in the clinical risk calculator requires access to the full training dataset as the user is using the calculator in order to allow SHAP to generate perturbations around a prediction to understand the local behaviors of the model. Involvement of training data in a web-based context represents a data privacy concern. To facilitate SHAP implementation, we will generate a synthetic dataset that mimics the distribution and characteristics of our training dataset for SHAP perturbations using a Differentially Private Conditional Tabular Generative Adversarial Network (DP-CTGAN) [48]. DP-CTGAN uses random noise during network training and data generation to offer a privacy guarantee. Hyperparameters of the DP-CTGAN will be tuned manually to minimize generator and discriminator loss. The privacy budget ε will be set to between 0 and 10. We aim to generate a synthetic dataset that yields similar SHAP value predictions and SHAP base values for use with the clinical risk calculator.

### Statistical softwares

The modeling and evaluation processes will be completed using Python 3.11. Boruta-based feature selection will be conducted using the *BorutaPy* package, and LASSO-based feature selection will be conducted using *scikit-learn*. ML models will be fitted, tuned, calibrated, and evaluated using *scikit-learn, lightgbm, xgboost, tensorflow, keras*, and *scikit-optimize*. Permutation importance will be assessed using *eli5*, and permutation SHAP will be implemented using *shap*.

### Patient and public involvement

None.

## ETHICS AND DISSEMINATION

The proposed study has received ethics approval from the Mayo Clinic Institutional Review Board (IRB #23-007416). The requirement for informed consent is waived by institutional review. The development process of the final model and performance measures yielded by nested CV will be described in a research article to be disseminated in a peer-reviewed academic journal.

## DISCUSSION

While traditional statistical modeling is useful for identifying predictors of hypertriglyceridemia in mechanically ventilated patients receiving continuous propofol infusion, it is difficult to systematically translate these findings to actual clinical practice. By leveraging ML-based modeling approaches and by incorporating ML models into an accessible clinical risk calculator, we aim to develop a tool that could guide clinicians in making informed decisions surrounding the choice of sedation regimens and potentially reduce the incidence of propofol-related hypertriglyceridemia and associated adverse events such as acute pancreatitis.

A notable use case for our proposed clinical risk calculator is to identify patients who may benefit from increased triglyceride level monitoring. In the absence of widely-accepted guidelines and protocols, routine measurements of triglycerides among patients receiving IMV with propofol sedation is often *ad hoc* and remains care-team dependent. Previous studies have shown that only 15-24% of intubated patients with propofol sedation receive routine measurements of triglyceride levels, with about a fifth of these patients having elevated triglyceride levels >400 mg/dL [5]. It is possible that a ML model can identify specific patient characteristics at the time of ICU admission or intubation that can predict patient risks of developing hypertriglyceridemia, so that triglyceride levels in these patients can be monitored and used to tailor the patients’ sedation regimen accordingly.

The proposed ML modeling study has several notable strengths. First, we propose the use of multiple well-established feature selection methods to reduce the dimensionality of our training dataset. This helps us to keep only the most important predictors of hypertriglyceridemia in our models, and to avoid the “curse of dimensionality”, including overfitting the models on noise in the data. Secondly, we aim to use robust external validation methods involving a nested CV framework with LOSO-CV. This approach allows the generalizability of our modeling process to be rigorously and repeatedly evaluated across different hospital settings, which improves the external validity of our performance metrics. Third, we aim to trial a diverse set of popular ML algorithms, including classical, ensemble, and neural network models, along with state-of-the-art hyperparameter tuning techniques using Bayesian optimization. This exhaustive approach will allow us to identify the most optimal ML model configuration for our dataset and target outcome. Lastly, we considered the clinical applicability of our ML models by proposing to integrate both global and local feature explainability to assess how our final model makes its predictions, as well as by integrating our final model into a web-based clinical risk calculator.

Despite these strengths, we foresee several potential challenges associated with the proposed study. First, the study relies on retrospective data, which can introduce biases relating to misclassification from potential errors in charting and missing data. The retrospective nature of the data also results in limited granularity on the exact severity of comorbid conditions such as liver disease and prevents us from capturing baseline triglyceride values which precludes their inclusion in the model. These shortcomings in our dataset highlights the need for additional prospective work to incorporate and further assess these features in future works. Secondly, while the proposed nested CV approach is thorough, it will be computationally intensive and time-consuming. This limits the feasibility of future replications of the proposed study methodology, especially in resource-constrained settings or when using larger datasets. Lastly, while the generalizability of our dataset is improved by including information from multiple Mayo Clinic sites, these sites may share similar treatment or data recording protocols that reduce the external validity of our developed models.

Overall, our protocol outlines a comprehensive approach to developing and validating a clinical tool designed to provide a personalized risk estimate of developing hypertriglyceridemia when sedated with continuous propofol infusion while on IMV. In the absence of routine triglyceride monitoring, it remains important to prevent the development of hypertriglyceridemia in an effort to avoid a sequela of adverse events.

## Data Availability

No additional data is available to support this protocol.

## Authors’ contributions

Jiawen Deng, Kiyan Heybati, and Hemang Yadav contributed equally to the conception and methodological design of the proposed study, and all three authors contributed equally to the drafting and review/editing of the protocol manuscript.

