## Supplemental Table for "Protocol for the development and validation of machine-learning models for predicting the risk of hypertriglyceridemia in critically ill patients receiving propofol sedation using retrospective data"

### Supplementary Material

**Table S1:** List of hyperparameters to be tuned for LASSO-penalized logistic regression, p.2
**Table S2:** List of hyperparameters to be tuned for ridge-penalized logistic regression, p.2
**Table S3:** List of hyperparameters to be tuned for elastic net-penalized logistic regression, p.3
**Table S4:** List of hyperparameters to be tuned for linear kernel SVM, p.4
**Table S5:** List of hyperparameters to be tuned for radial basis function kernel SVM, p.4
**Table S6:** List of hyperparameters to be tuned for polynomial kernel SVM, p.5
**Table S7:** List of hyperparameters to be tuned for sigmoid kernel SVM, p.6
**Table S8:** List of hyperparameters to be tuned for random decision forest, p.7
**Table S9:** List of hyperparameters to be tuned for LightGBM with imputation and GBDT, p.8
**Table S10:** List of hyperparameters to be tuned for LightGBM with imputation and DART, p.10
**Table S11:** List of hyperparameters to be tuned for LightGBM without imputation and GBDT, p.11
**Table S12:** List of hyperparameters to be tuned for LightGBM without imputation and DART, p.12
**Table S13:** List of hyperparameters to be tuned for XGBoost with imputation and GBDT, p.14
**Table S14:** List of hyperparameters to be tuned for XGBoost with imputation and DART, p.15
**Table S15:** List of hyperparameters to be tuned for XGBoost without imputation and GBDT, p.17
**Table S16:** List of hyperparameters to be tuned for XGBoost without imputation and DART, p.18
**Table S17:** List of hyperparameters to be tuned for neural network models, p.20

**Table S1:** List of hyperparameters to be tuned for LASSO-penalized logistic regression

| Hyperparameter | Hyperparameter description | Hyperparameter search space |
|---|---|---|
| *penalty* | Specifies the type of regularization used to prevent overfitting by penalizing large coefficient values. | Categorical(['l1']) |
| *C* | Controls the regularization strength. It is the inverse of the regularization parameter. Smaller values of *C* imply stronger regularization, while larger values of *C* decrease the regularization effect. | Real(1e-4, 1e4, prior='log-uniform') |
| *solver* | Specifies the algorithm used to optimize the model's coefficients during training. | Categorical(['liblinear','saga']) |
| *class_weight* | Used to handle imbalanced datasets by assigning different weights to classes. When set to "balanced," it automatically adjusts weights inversely proportional to class frequencies in the input data. | Categorical([None, 'balanced']) |
| *pca* | Controls whether PCA is applied. | Categorical([PCA(random_state=SEED, svd_solver="full", n_components=0.95), 'passthrough']) |

**Table S2:** List of hyperparameters to be tuned for ridge-penalized logistic regression

| Hyperparameter | Hyperparameter description | Hyperparameter search space |
|---|---|---|
| *penalty* | Specifies the type of regularization used to prevent overfitting by penalizing large coefficient values. | Categorical(['l2']) |
| *C* | Controls the regularization strength. It is the inverse of the regularization parameter. Smaller values of *C* imply stronger regularization, while larger values of *C* decrease the regularization effect. | Real(1e-4, 1e4, prior='log-uniform') |

| | | |
|---|---|---|
| *solver* | Specifies the algorithm used to optimize the model's coefficients during training. | Categorical(['lbfgs','liblinear','newton-cg','newton-cholesky','saga','sag']) |
| *class_weight* | Used to handle imbalanced datasets by assigning different weights to classes. When set to "balanced," it automatically adjusts weights inversely proportional to class frequencies in the input data. | Categorical([None, 'balanced']) |
| *pca* | Controls whether PCA is applied. | Categorical([PCA(random_state=SEED, svd_solver="full", n_components=0.95), 'passthrough']) |

**Table S3:** List of hyperparameters to be tuned for elastic net-penalized logistic regression

| Hyperparameter | Hyperparameter description | Hyperparameter search space |
|---|---|---|
| *penalty* | Specifies the type of regularization used to prevent overfitting by penalizing large coefficient values. | Categorical(['elasticnet']) |
| *C* | Controls the regularization strength. It is the inverse of the regularization parameter. Smaller values of *C* imply stronger regularization, while larger values of *C* decrease the regularization effect. | Real(1e-4, 1e4, prior='log-uniform') |
| *solver* | Specifies the algorithm used to optimize the model's coefficients during training. | Categorical(['saga']) |
| *l1_ratio* | Controls the balance between L1 and L2 regularization. | Real(0, 1, prior='uniform') |
| *class_weight* | Used to handle imbalanced datasets by assigning different weights to classes. When set to "balanced," it automatically adjusts weights inversely proportional to class frequencies in the input data. | Categorical([None, 'balanced']) |

| | | |
|---|---|---|
| *pca* | Controls whether PCA is applied. | Categorical([PCA(random_state=SEED, svd_solver="full", n_components=0.95), 'passthrough']) |

**Table S4:** List of hyperparameters to be tuned for linear kernel SVM

| Hyperparameter | Hyperparameter description | Hyperparameter search space |
|---|---|---|
| *C* | Controls the trade-off between maximizing the margin and minimizing the classification error. A small value of *C* allows the classifier to have a larger margin at the cost of misclassifying more points. A large value of *C* prioritizes correctly classifying all training examples. | Real(1e-4, 1e4, prior='log-uniform') |
| *kernel* | Specifies the type of kernel function used to transform the data into a higher-dimensional space. | Categorical(['linear']) |
| *class_weight* | Used to handle imbalanced datasets by assigning different weights to classes. When set to "balanced," it automatically adjusts weights inversely proportional to class frequencies in the input data. | Categorical([None, 'balanced']) |
| *pca* | Controls whether PCA is applied. | Categorical([PCA(random_state=SEED, svd_solver="full", n_components=0.95), 'passthrough']) |

**Table S5:** List of hyperparameters to be tuned for radial basis function kernel SVM

| Hyperparameter | Hyperparameter description | Hyperparameter search space |
|---|---|---|
| *C* | Controls the trade-off between maximizing | Real(1e-4, 1e4, prior='log- |

| | | |
|---|---|---|
| | the margin and minimizing the classification error. A small value of *C* allows the classifier to have a larger margin at the cost of misclassifying more points. A large value of *C* prioritizes correctly classifying all training examples. | uniform') |
| *kernel* | Specifies the type of kernel function used to transform the data into a higher-dimensional space. | Categorical(['rbf']) |
| *gamma* | Controls the curvature of the decision boundary. | Real(1e-4, 1e4, prior='log-uniform') |
| *class_weight* | Used to handle imbalanced datasets by assigning different weights to classes. When set to "balanced," it automatically adjusts weights inversely proportional to class frequencies in the input data. | Categorical([None, 'balanced']) |
| *pca* | Controls whether PCA is applied. | Categorical([PCA(random_state=SEED, svd_solver="full", n_components=0.95), 'passthrough']) |

**Table S6:** List of hyperparameters to be tuned for polynomial kernel SVM

| **Hyperparameter** | **Hyperparameter description** | **Hyperparameter search space** |
|---|---|---|
| *C* | Controls the trade-off between maximizing the margin and minimizing the classification error. A small value of *C* allows the classifier to have a larger margin at the cost of misclassifying more points. A large value of *C* prioritizes correctly classifying all training examples. | Real(1e-4, 1e4, prior='log-uniform') |
| *kernel* | Specifies the type of kernel function used to transform the data into a higher-dimensional space. | Categorical(['poly']) |

| | | |
|---|---|---|
| *gamma* | Controls the curvature of the decision boundary. | Real(1e-4, 1e4, prior='log-uniform') |
| *degree* | Specifies the degree of the polynomial kernel function and controls the complexity of the decision boundary. | Categorical([2, 3, 4, 5]) |
| *coef0* | Controls the influence of the independent term in the kernel function. Adds a bias to the decision boundary, which allows the model to fit the data more flexibly. | Real(-15, 15, prior='uniform') |
| *class_weight* | Used to handle imbalanced datasets by assigning different weights to classes. When set to "balanced," it automatically adjusts weights inversely proportional to class frequencies in the input data. | Categorical([None, 'balanced']) |
| *pca* | Controls whether PCA is applied. | Categorical([PCA(random_state=SEED, svd_solver="full", n_components=0.95), 'passthrough']) |

**Table S7:** List of hyperparameters to be tuned for sigmoid kernel SVM

| Hyperparameter | Hyperparameter description | Hyperparameter search space |
|---|---|---|
| *C* | Controls the trade-off between maximizing the margin and minimizing the classification error. A small value of *C* allows the classifier to have a larger margin at the cost of misclassifying more points. A large value of *C* prioritizes correctly classifying all training examples. | Real(1e-4, 1e4, prior='log-uniform') |
| *kernel* | Specifies the type of kernel function used to transform the data into a higher-dimensional space. | Categorical(['sigmoid']) |
| *gamma* | Controls the curvature of the decision boundary. | Real(1e-4, 1e4, prior='log-uniform') |

| | | |
|---|---|---|
| *coef0* | Controls the influence of the independent term in the kernel function. Adds a bias to the decision boundary, which allows the model to fit the data more flexibly. | Real(-15, 15, prior='uniform') |
| *class_weight* | Used to handle imbalanced datasets by assigning different weights to classes. When set to "balanced," it automatically adjusts weights inversely proportional to class frequencies in the input data. | Categorical([None, 'balanced']) |
| *pca* | Controls whether PCA is applied. | Categorical([PCA(random_state=SEED, svd_solver="full", n_components=0.95), 'passthrough']) |

**Table S8:** List of hyperparameters to be tuned for random decision forest

| Hyperparameter | Hyperparameter description | Hyperparameter search space |
|---|---|---|
| *n_estimators* | Specifies the number of decision trees in the forest and controls the size of the ensemble model. | Integer(2, 2000) |
| *criterion* | Specifies the function used to measure the quality of a split during the construction of decision trees within the forest. | Categorical(['gini', 'entropy', 'log_loss']) |
| *max_depth* | Controls the maximum depth of each individual decision tree in the forest. | Integer(2, 100) |
| *min_samples_split* | Controls the minimum number of samples required to split an internal node. | Real(0.00, 0.50, prior='uniform') |
| *min_samples_leaf* | Specifies the minimum number of samples required to be at a leaf node. | Real(0.00, 0.50, prior='uniform') |
| *min_weight_fraction_leaf* | Specifies the minimum weighted fraction of the total sum of weights required to be at a leaf node. | Real(0.00, 0.50, prior='uniform') |

| | | |
|---|---|---|
| *max_features* | Determines the maximum number of features considered when splitting a node in each decision tree. | Real(0.01, 0.99, prior='uniform') |
| *max_leaf_nodes* | Controls the maximum number of leaf nodes in each decision tree within the forest. | Integer(2, 1000) |
| *min_impurity_decrease* | Specifies the minimum decrease in impurity required for a node to be split. | Real(0.0, 0.1, prior='uniform') |
| *class_weight* | Used to handle imbalanced datasets by assigning different weights to classes. When set to "balanced," it automatically adjusts weights inversely proportional to class frequencies in the input data. When set to "balanced_subsample", it adjusts the class weights for each individual decision tree within the forest, based on the class distribution of the bootstrap sample used to train that tree. | Categorical(['balanced', 'balanced_subsample', None]) |
| *max_samples* | Specifies the number or proportion of samples to draw from the training data to train each individual decision tree in the forest. | Real(0.01, 1.00, prior='uniform') |
| *ccp_alpha* | Sets a threshold value for the cost-complexity pruning process, which prunes the branches of each decision tree to prevent overfitting. | Real(0.0, 0.1, prior='uniform') |
| *pca* | Controls whether PCA is applied. | Categorical([PCA(random_state=SEED, svd_solver="full", n_components=0.95), 'passthrough']) |

**Table S9:** List of hyperparameters to be tuned for LightGBM with imputation and GBDT

| Hyperparameter | Hyperparameter description | Hyperparameter search space |
|---|---|---|
| *num_leaves* | Specifies the maximum number of leaves in | Integer(2, 1000) |

| | | |
|---|---|---|
| | one tree. | |
| *max_depth* | Limits the maximum depth of the tree. | Integer(2, 100) |
| *learning_rate* | Controls the contribution of each tree to the final model. | Real(0.0001, 0.5, prior='log-uniform') |
| *n_estimators* | Determines the number of boosting iterations/the number of trees. | Integer(2, 2000) |
| *is_unbalance* | When set to "true", adjusts the class weights automatically to address imbalanced datasets. | Categorical([True, False]) |
| *min_split_gain* | The minimum gain required to make a further partition on a leaf node of the tree. Higher values lead to more conservative splits. | Real(0.0, 1.0, prior='uniform') |
| *min_child_samples* | The minimum number of data points needed in a leaf. Prevents the model from learning overly specific patterns by requiring a minimum number of samples to make a split. | Integer(2, 5000) |
| *subsample* | The fraction of samples used for training each tree. | Real(0.01, 1.00, prior='uniform') |
| *subsample_freq* | The frequency of applying subsampling. | Integer(-1, 100) |
| *colsample_bytree* | The fraction of features to be randomly selected for each tree. | Real(0.01, 1.00, prior='uniform') |
| *reg_alpha* | L1 regularization term on weights to encourage sparsity (fewer features). | Real(0.0001, 1000, prior='log-uniform') |
| *reg_lambda* | L2 regularization term on weights to discourage complex models. | Real(0.0001, 1000, prior='log-uniform') |
| *boosting_type* | Specifies the type of boosting algorithm to use. | Categorical(['gbdt']) |
| *pca* | Controls whether PCA is applied. | Categorical([PCA(random_state=SEED, svd_solver="full", n_components=0.95), 'passthrough']) |

**Table S10:** List of hyperparameters to be tuned for LightGBM with imputation and DART

| Hyperparameter | Hyperparameter description | Hyperparameter search space |
|---|---|---|
| *num_leaves* | Specifies the maximum number of leaves in one tree. | Integer(2, 1000) |
| *max_depth* | Limits the maximum depth of the tree. | Integer(2, 100) |
| *learning_rate* | Controls the contribution of each tree to the final model. | Real(0.0001, 0.5, prior='log-uniform') |
| *n_estimators* | Determines the number of boosting iterations/the number of trees. | Integer(2, 2000) |
| *is_unbalance* | When set to "true", adjusts the class weights automatically to address imbalanced datasets. | Categorical([True, False]) |
| *min_split_gain* | The minimum gain required to make a further partition on a leaf node of the tree. Higher values lead to more conservative splits. | Real(0.0, 1.0, prior='uniform') |
| *min_child_samples* | The minimum number of data points needed in a leaf. Prevents the model from learning overly specific patterns by requiring a minimum number of samples to make a split. | Integer(2, 5000) |
| *subsample* | The fraction of samples used for training each tree. | Real(0.01, 1.00, prior='uniform') |
| *subsample_freq* | The frequency of applying subsampling. | Integer(-1, 100) |
| *colsample_bytree* | The fraction of features to be randomly selected for each tree. | Real(0.01, 1.00, prior='uniform') |
| *reg_alpha* | L1 regularization term on weights to encourage sparsity (fewer features). | Real(0.0001, 1000, prior='log-uniform') |
| *reg_lambda* | L2 regularization term on weights to discourage complex models. | Real(0.0001, 1000, prior='log-uniform') |

| | | |
|---|---|---|
| *uniform_drop* | When set to True, ensures that trees are uniformly dropped during the training process, rather than being dropped randomly based on their weights. | Categorical([True, False]) |
| *xgboost_dart_mode* | When set to True, changes the behavior of the DART boosting method to be more similar to XGBoost's implementation of DART. | Categorical([True, False]) |
| *drop_rate* | Defines the probability of dropping each tree during the boosting process in DART. | Real(0, 1, prior='uniform') |
| *skip_drop* | Sets the probability of skipping the dropout procedure during a boosting iteration. | Real(0, 1, prior='uniform') |
| *max_drop* | Specifies the maximum number of trees that can be dropped during the boosting process in each iteration. | Integer(0, 100) |
| *drop_seed* | Sets the random seed for the dropout procedure, ensuring reproducibility of results. | Categorical([SEED]) |
| *boosting_type* | Specifies the type of boosting algorithm to use. | Categorical(['dart']) |
| *pca* | Controls whether PCA is applied. | Categorical([PCA(random_state=SEED, svd_solver="full", n_components=0.95), 'passthrough']) |

**Table S11:** List of hyperparameters to be tuned for LightGBM without imputation and GBDT

| Hyperparameter | Hyperparameter description | Hyperparameter search space |
|---|---|---|
| *num_leaves* | Specifies the maximum number of leaves in one tree. | Integer(2, 1000) |
| *max_depth* | Limits the maximum depth of the tree. | Integer(2, 100) |
| *learning_rate* | Controls the contribution of each tree to the | Real(0.0001, 0.5, |

| | | |
|---|---|---|
| | final model. | prior='log-uniform') |
| *n_estimators* | Determines the number of boosting iterations/the number of trees. | Integer(2, 2000) |
| *is_unbalance* | When set to “true”, adjusts the class weights automatically to address imbalanced datasets. | Categorical([True, False]) |
| *min_split_gain* | The minimum gain required to make a further partition on a leaf node of the tree. Higher values lead to more conservative splits. | Real(0.0, 1.0, prior='uniform') |
| *min_child_samples* | The minimum number of data points needed in a leaf. Prevents the model from learning overly specific patterns by requiring a minimum number of samples to make a split. | Integer(2, 5000) |
| *subsample* | The fraction of samples used for training each tree. | Real(0.01, 1.00, prior='uniform') |
| *subsample_freq* | The frequency of applying subsampling. | Integer(-1, 100) |
| *colsample_bytree* | The fraction of features to be randomly selected for each tree. | Real(0.01, 1.00, prior='uniform') |
| *reg_alpha* | L1 regularization term on weights to encourage sparsity (fewer features). | Real(0.0001, 1000, prior='log-uniform') |
| *reg_lambda* | L2 regularization term on weights to discourage complex models. | Real(0.0001, 1000, prior='log-uniform') |
| *boosting_type* | Specifies the type of boosting algorithm to use. | Categorical(['gbdt']) |
| *impute* | Controls whether imputation is applied. | Categorical(['passthrough']) |
| *pca* | Controls whether PCA is applied. | Categorical(['passthrough']) |

**Table S12:** List of hyperparameters to be tuned for LightGBM without imputation and DART

| Hyperparameter | Hyperparameter description | Hyperparameter search space |
|---|---|---|
| *num_leaves* | Specifies the maximum number of leaves in one tree. | Integer(2, 1000) |
| *max_depth* | Limits the maximum depth of the tree. | Integer(2, 100) |
| *learning_rate* | Controls the contribution of each tree to the final model. | Real(0.0001, 0.5, prior='log-uniform') |
| *n_estimators* | Determines the number of boosting iterations/the number of trees. | Integer(2, 2000) |
| *is_unbalance* | When set to "true", adjusts the class weights automatically to address imbalanced datasets. | Categorical([True, False]) |
| *min_split_gain* | The minimum gain required to make a further partition on a leaf node of the tree. Higher values lead to more conservative splits. | Real(0.0, 1.0, prior='uniform') |
| *min_child_samples* | The minimum number of data points needed in a leaf. Prevents the model from learning overly specific patterns by requiring a minimum number of samples to make a split. | Integer(2, 5000) |
| *subsample* | The fraction of samples used for training each tree. | Real(0.01, 1.00, prior='uniform') |
| *subsample_freq* | The frequency of applying subsampling. | Integer(-1, 100) |
| *colsample_bytree* | The fraction of features to be randomly selected for each tree. | Real(0.01, 1.00, prior='uniform') |
| *reg_alpha* | L1 regularization term on weights to encourage sparsity (fewer features). | Real(0.0001, 1000, prior='log-uniform') |
| *reg_lambda* | L2 regularization term on weights to discourage complex models. | Real(0.0001, 1000, prior='log-uniform') |
| *uniform_drop* | When set to True, ensures that trees are uniformly dropped during the training process, rather than being dropped randomly based on their weights. | Categorical([True, False]) |

| | | |
|---|---|---|
| *xgboost_dart_mode* | When set to True, changes the behavior of the DART boosting method to be more similar to XGBoost's implementation of DART. | Categorical([True, False]) |
| *drop_rate* | Defines the probability of dropping each tree during the boosting process in DART. | Real(0, 1, prior='uniform') |
| *skip_drop* | Sets the probability of skipping the dropout procedure during a boosting iteration. | Real(0, 1, prior='uniform') |
| *max_drop* | Specifies the maximum number of trees that can be dropped during the boosting process in each iteration. | Integer(0, 100) |
| *drop_seed* | Sets the random seed for the dropout procedure, ensuring reproducibility of results. | Categorical([SEED]) |
| *boosting_type* | Specifies the type of boosting algorithm to use. | Categorical(['dart']) |
| *impute* | Controls whether imputation is applied. | Categorical(['passthrough']) |
| *pca* | Controls whether PCA is applied. | Categorical(['passthrough']) |

**Table S13:** List of hyperparameters to be tuned for XGBoost with imputation and GBDT

| Hyperparameter | Hyperparameter description | Hyperparameter search space |
|---|---|---|
| *n_estimators* | The number of trees to be built in the model. | Integer(2, 2000) |
| *max_depth* | The maximum depth of each tree. | Integer(2, 100) |
| *max_leaves* | The maximum number of leaves or terminal nodes in each tree. | Integer(2, 1000) |
| *grow_policy* | Determines the tree construction strategy, either depth-wise (traditional) or loss-guided, which prioritizes branches with greater potential for reducing loss. | Categorical(['depthwise', 'lossguide']) |

| | | |
|---|---|---|
| *learning_rate* | The step size shrinkage used to prevent overfitting by controlling the contribution of each tree. | Real(0.0001, 0.5, prior='log-uniform') |
| *booster* | Specifies the type of boosting algorithm to use. | Categorical(['gbtree']) |
| *tree_method* | The algorithm used for constructing trees. | Categorical(['approx', 'hist', 'auto']) |
| *gamma* | The minimum loss reduction required for a split to be made. | Real(0.0001, 1000, prior='log-uniform') |
| *min_child_weight* | The minimum sum of instance weight (hessian) needed in a child. | Integer(0, 1000) |
| *max_delta_step* | The maximum step size allowed for each tree's weight. | Real(0, 10, prior='uniform') |
| *subsample* | The fraction of training data used to grow each tree. | Real(0.01, 1, prior='uniform') |
| *colsample_bytree* | The fraction of features to be randomly sampled for each tree. | Real(0.01, 1) |
| *colsample_bylevel* | The fraction of features to be sampled for each level of the tree. | Real(0.01, 1) |
| *colsample_bynode* | The fraction of features to be sampled for each node split. | Real(0.01, 1) |
| *scale_pos_weight* | The scaling factor for positive class weights. | Real(0.01, 100, prior='log-uniform') |
| *reg_alpha* | The L1 regularization term on weights. | Real(0.0001, 1000, prior='log-uniform') |
| *reg_lambda* | The L2 regularization term on weights. | Real(0.0001, 1000, prior='log-uniform') |
| *pca* | Controls whether PCA is applied. | Categorical([PCA(random_state=SEED, svd_solver="full", n_components=0.95), 'passthrough']) |

**Table S14:** List of hyperparameters to be tuned for XGBoost with imputation and DART

| Hyperparameter | Hyperparameter description | Hyperparameter search space |
|---|---|---|
| *n_estimators* | The number of trees to be built in the model. | Integer(2, 2000) |
| *max_depth* | The maximum depth of each tree. | Integer(2, 100) |
| *max_leaves* | The maximum number of leaves or terminal nodes in each tree. | Integer(2, 1000) |
| *grow_policy* | Determines the tree construction strategy, either depth-wise (traditional) or loss-guided, which prioritizes branches with greater potential for reducing loss. | Categorical(['depthwise', 'lossguide']) |
| *learning_rate* | The step size shrinkage used to prevent overfitting by controlling the contribution of each tree. | Real(0.0001, 0.5, prior='log-uniform') |
| *booster* | Specifies the type of boosting algorithm to use. | Categorical(['dart']) |
| *tree_method* | The algorithm used for constructing trees. | Categorical(['approx', 'hist', 'auto']) |
| *gamma* | The minimum loss reduction required for a split to be made. | Real(0.0001, 1000, prior='log-uniform') |
| *min_child_weight* | The minimum sum of instance weight (hessian) needed in a child. | Integer(0, 1000) |
| *max_delta_step* | The maximum step size allowed for each tree's weight. | Real(0, 10, prior='uniform') |
| *subsample* | The fraction of training data used to grow each tree. | Real(0.01, 1, prior='uniform') |
| *colsample_bytree* | The fraction of features to be randomly sampled for each tree. | Real(0.01, 1) |
| *colsample_bylevel* | The fraction of features to be sampled for each level of the tree. | Real(0.01, 1) |
| *colsample_bynode* | The fraction of features to be sampled for each node split. | Real(0.01, 1) |

| | | |
|---|---|---|
| *scale_pos_weight* | The scaling factor for positive class weights. | Real(0.01, 100, prior='log-uniform') |
| *reg_alpha* | The L1 regularization term on weights. | Real(0.0001, 1000, prior='log-uniform') |
| *reg_lambda* | The L2 regularization term on weights. | Real(0.0001, 1000, prior='log-uniform') |
| *sample_type* | Specifies the type of sampling method used during boosting when using the 'dart' booster. | Categorical(['uniform', 'weighted']) |
| *normalize_type* | Defines the normalization method used when using the 'dart' booster. | Categorical(['tree', 'forest']) |
| *rate_drop* | Sets the dropout rate for the 'dart' booster. | Real(0.00, 1, prior='uniform') |
| *skip_drop* | Controls the probability of skipping the dropout process in the 'dart' booster. | Real(0.00, 1, prior='uniform') |
| *pca* | Controls whether PCA is applied. | Categorical([PCA(random_state=SEED, svd_solver="full", n_components=0.95), 'passthrough']) |

**Table S15:** List of hyperparameters to be tuned for XGBoost without imputation and GBDT

| **Hyperparameter** | **Hyperparameter description** | **Hyperparameter search space** |
|---|---|---|
| *n_estimators* | The number of trees to be built in the model. | Integer(2, 2000) |
| *max_depth* | The maximum depth of each tree. | Integer(2, 100) |
| *max_leaves* | The maximum number of leaves or terminal nodes in each tree. | Integer(2, 1000) |
| *grow_policy* | Determines the tree construction strategy, either depth-wise (traditional) or loss-guided, which prioritizes branches with greater potential for reducing loss. | Categorical(['depthwise', 'lossguide']) |

| | | |
|---|---|---|
| *learning_rate* | The step size shrinkage used to prevent overfitting by controlling the contribution of each tree. | Real(0.0001, 0.5, prior='log-uniform') |
| *booster* | Specifies the type of boosting algorithm to use. | Categorical(['gbtree']) |
| *tree_method* | The algorithm used for constructing trees. | Categorical(['approx', 'hist', 'auto']) |
| *gamma* | The minimum loss reduction required for a split to be made. | Real(0.0001, 1000, prior='log-uniform') |
| *min_child_weight* | The minimum sum of instance weight (hessian) needed in a child. | Integer(0, 1000) |
| *max_delta_step* | The maximum step size allowed for each tree's weight. | Real(0, 10, prior='uniform') |
| *subsample* | The fraction of training data used to grow each tree. | Real(0.01, 1, prior='uniform') |
| *colsample_bytree* | The fraction of features to be randomly sampled for each tree. | Real(0.01, 1) |
| *colsample_bylevel* | The fraction of features to be sampled for each level of the tree. | Real(0.01, 1) |
| *colsample_bynode* | The fraction of features to be sampled for each node split. | Real(0.01, 1) |
| *scale_pos_weight* | The scaling factor for positive class weights. | Real(0.01, 100, prior='log-uniform') |
| *reg_alpha* | The L1 regularization term on weights. | Real(0.0001, 1000, prior='log-uniform') |
| *reg_lambda* | The L2 regularization term on weights. | Real(0.0001, 1000, prior='log-uniform') |
| *impute* | Controls whether imputation is applied. | Categorical(['passthrough']) |
| *pca* | Controls whether PCA is applied. | Categorical(['passthrough']) |

**Table S16:** List of hyperparameters to be tuned for XGBoost without imputation and DART

| Hyperparameter | Hyperparameter description | Hyperparameter search space |
|---|---|---|
| *n_estimators* | The number of trees to be built in the model. | Integer(2, 2000) |
| *max_depth* | The maximum depth of each tree. | Integer(2, 100) |
| *max_leaves* | The maximum number of leaves or terminal nodes in each tree. | Integer(2, 1000) |
| *grow_policy* | Determines the tree construction strategy, either depth-wise (traditional) or loss-guided, which prioritizes branches with greater potential for reducing loss. | Categorical(['depthwise', 'lossguide']) |
| *learning_rate* | The step size shrinkage used to prevent overfitting by controlling the contribution of each tree. | Real(0.0001, 0.5, prior='log-uniform') |
| *booster* | Specifies the type of boosting algorithm to use. | Categorical(['dart']) |
| *tree_method* | The algorithm used for constructing trees. | Categorical(['approx', 'hist', 'auto']) |
| *gamma* | The minimum loss reduction required for a split to be made. | Real(0.0001, 1000, prior='log-uniform') |
| *min_child_weight* | The minimum sum of instance weight (hessian) needed in a child. | Integer(0, 1000) |
| *max_delta_step* | The maximum step size allowed for each tree's weight. | Real(0, 10, prior='uniform') |
| *subsample* | The fraction of training data used to grow each tree. | Real(0.01, 1, prior='uniform') |
| *colsample_bytree* | The fraction of features to be randomly sampled for each tree. | Real(0.01, 1) |
| *colsample_bylevel* | The fraction of features to be sampled for each level of the tree. | Real(0.01, 1) |
| *colsample_bynode* | The fraction of features to be sampled for each node split. | Real(0.01, 1) |

| | | |
|---|---|---|
| *scale_pos_weight* | The scaling factor for positive class weights. | Real(0.01, 100, prior='log-uniform') |
| *reg_alpha* | The L1 regularization term on weights. | Real(0.0001, 1000, prior='log-uniform') |
| *reg_lambda* | The L2 regularization term on weights. | Real(0.0001, 1000, prior='log-uniform') |
| *sample_type* | Specifies the type of sampling method used during boosting when using the 'dart' booster. | Categorical(['uniform', 'weighted']) |
| *normalize_type* | Defines the normalization method used when using the 'dart' booster. | Categorical(['tree', 'forest']) |
| *rate_drop* | Sets the dropout rate for the 'dart' booster. | Real(0.00, 1, prior='uniform') |
| *skip_drop* | Controls the probability of skipping the dropout process in the 'dart' booster. | Real(0.00, 1, prior='uniform') |
| *impute* | Controls whether imputation is applied. | Categorical(['passthrough']) |
| *pca* | Controls whether PCA is applied. | Categorical(['passthrough']) |

**Table S17:** List of hyperparameters to be tuned for neural network models

| **Hyperparameter** | **Hyperparameter description** | **Hyperparameter search space** |
|---|---|---|
| *learning_rate* | Sets the learning rate for the AdamW optimizer. | Real(1e-6, 1e-1, prior='log-uniform') |
| *weight_decay* | Sets the weight decay rate for the AdamW optimizer. | Real(1e-6, 1e-1, prior='log-uniform') |
| *batch_size* | Sets the batch size used for each iteration of the model update. | Integer(4, 1024) |
| *n_layers* | Sets the number of hidden ReLU layers in the network. | Categorical([2, 3, 4]) |

| | | |
|---|---|---|
| *n_neurons* | Sets the number of neurons within each hidden ReLU layer in the network. | Integer(4, 1024) |
| *epochs* | Sets the number of epochs for the network training process. | Integer(1, 2000) |
| *dropout_#* | Sets the probability of the dropout layers. There will be one dropout hyperparameter for each dropout layer included in the network (depending on the n_layers parameter). | Real(0.01, 0.99, prior='uniform') |
| *pca* | Controls whether PCA is applied. | Categorical([PCA(random_state=SEED, svd_solver="full", n_components=0.95), 'passthrough']) |
